# Large Language Model Uncertainty Measurement and Calibration for Medical Diagnosis and Treatment

**DOI:** 10.1101/2024.06.06.24308399

**Authors:** Thomas Savage, John Wang, Robert Gallo, Abdessalem Boukil, Vishwesh Patel, Seyed Amir Ahmad Safavi-Naini, Ali Soroush, Jonathan H Chen

## Abstract

**Introduction:** The inability of Large Language Models (LLMs) to communicate uncertainty is a significant barrier to their use in medicine. Before LLMs can be integrated into patient care, the field must assess methods to measure uncertainty in ways that are useful to physician-users.

**Objective:** Evaluate the ability for uncertainty metrics to quantify LLM confidence when performing diagnosis and treatment selection tasks by assessing the properties of discrimination and calibration.

**Methods:** We examined Confidence Elicitation, Token Level Probability, and Sample Consistency metrics across GPT3.5, GPT4, Llama2 and Llama3. Uncertainty metrics were evaluated against three datasets of open-ended patient scenarios.

**Results:** Sample Consistency methods outperformed Token Level Probability and Confidence Elicitation methods. Sample Consistency by Sentence Embedding achieved the highest discrimination performance (ROC AUC 0.68–0.79) with poor calibration, while Sample Consistency by GPT Annotation achieved the second-best discrimination (ROC AUC 0.66-0.74) with more accurate calibration. Nearly all uncertainty metrics had better discriminative performance with diagnosis rather than treatment selection questions. Furthermore, verbalized confidence (Confidence Elicitation) was found to consistently over-estimate model confidence.

**Conclusions:** Sample Consistency is the most effective method for estimating LLM uncertainty of the metrics evaluated. Sample Consistency by Sentence Embedding can effectively estimate uncertainty if the user has a set of reference cases with which to re-calibrate their results, while Sample Consistency by GPT Annotation is more effective method if the user does not have reference cases and requires accurate raw calibration. Our results confirm LLMs are consistently over-confident when verbalizing their confidence through Confidence Elicitation.

## Introduction

Large language models (LLMs) have garnered significant interest from the medical community because of their abilities to analyze text data and generate useful responses.^1–4^ LLMs demonstrate impressive question-answering accuracy that rivals physicians for key tasks such as diagnosis and treatment selection.^5,6^ Despite this potential, a major obstacle to the use of language models in patient care is their difficulty expressing uncertainty.^7^

In medicine, assessing the uncertainty of a proposed diagnosis or treatment is integral for proper patient care. The clinician must decide whether there is sufficient confidence in an answer to make a definitive management decision such as prescribe a medicine or recommend surgery which have associated risks. LLMs use probabilities to generate text, but they may state answers confidently even if the underlying probability of its response has substantial uncertainty. Therefore, before LLMs are integrated into patient care, the field must assess methods to measure LLM response uncertainty in a way that is useful to physician-users.

Uncertainty measurement is similarly important for the development of LLM applications that use Retrieval Augmented Generation (RAG). RAG is a popular technique to increase LLM accuracy and reduce hallucinations when a language model is asked a question outside the scope of its training data. RAG searches a library for any text that can be used as helpful context to better answer the question outside of its base knowledge scope.^8^ A key challenge of modern LLM systems is selectively deploying RAG only when a model is uncertain of an answer because extra context provided by RAG can errantly lead the model into an incorrect response.^9^ A standard best practice for assessing model uncertainty is not yet established,^10,11^ in turn evaluation of methods to estimate LLM uncertainty are important for building medical LLM-RAG systems.

In this study we compare three methods of uncertainty measurement: Confidence Elicitation, Token-Level Probabilities, and Sample Consistency among large language models GPT 3.5^12^, GPT 4^13^, Llama2-70B^14^ and Llama3-70B^15^ for both their discrimination and calibration properties in estimating model uncertainty for the clinical reasoning tasks of medical diagnosis and treatment selection

## Background

### Discrimination and Calibration

Uncertainty metrics are evaluated from the perspective of two properties: Discrimination and Calibration. Discrimination is the ability of an uncertainty measure to differentiate between correct and incorrect answers, essentially how accurate the metric is in determining whether the LLM is correct for a given question.^16^ In medicine, discrimination is often measured by odds ratio. In computer science, uncertainty discrimination is often measured by the Receiver Operating Characteristic Area Under the Curve (ROC AUC)^17–19^ that summarizes the tradeoff between sensitivity and specificity, acknowledging that uncertainty measures are a surrogate for true uncertainty probability. Calibration goes a step further and assesses whether the uncertainty metric’s predicted probability of correctness agrees with the observed probability. Calibration can be evaluated by a calibration plot as well as Expected Calibration Error and Brier’s Score.^20^

Frequently an uncertainty metric can demonstrate strong discrimination but poor calibration, which drastically reduces the utility for a clinician. For example, a model may give higher certainty to correct responses compared to incorrect responses but is overconfident in its uncertainty measure across responses. If the uncertainty metric suggests 99% confidence that the response is correct, but the observed accuracy is only 80%, this may lead a clinician to make inappropriate decisions, such as prematurely concluding a patient has a specific diagnosis when there is still a sizable chance of other diagnoses.^21^ A useful prediction metric should demonstrate both strong discrimination as well as accurate calibration in order to be clinically useful.

### Uncertainty Metrics

Our work builds on existing literature of LLM uncertainty measures in the fields of arithmetic, logic and symbolic reasoning.^17,19,22,23^ Methods of uncertainty determination are split into three categories: Deterministic, Sample Consistency and Ensemble methods (Figure 1). Deterministic methods are uncertainty measures that are calculated from one forward pass of a model. Sample Consistency methods are measures calculated from multiple forward passes of a single model, comparing similarity of responses (called consistency) to estimate uncertainty. Finally, Ensemble methods are measures calculated from a collection of single forward passes from multiple models, also comparing similarity of responses to estimate uncertainty. Ensemble methods are expensive in terms of computation time and cost, which are especially compounded for medical applications that require HIPAA level security. As a result, Ensemble methods are impractical for clinical use and thus will not be included in our investigation.

**Figure 1.**
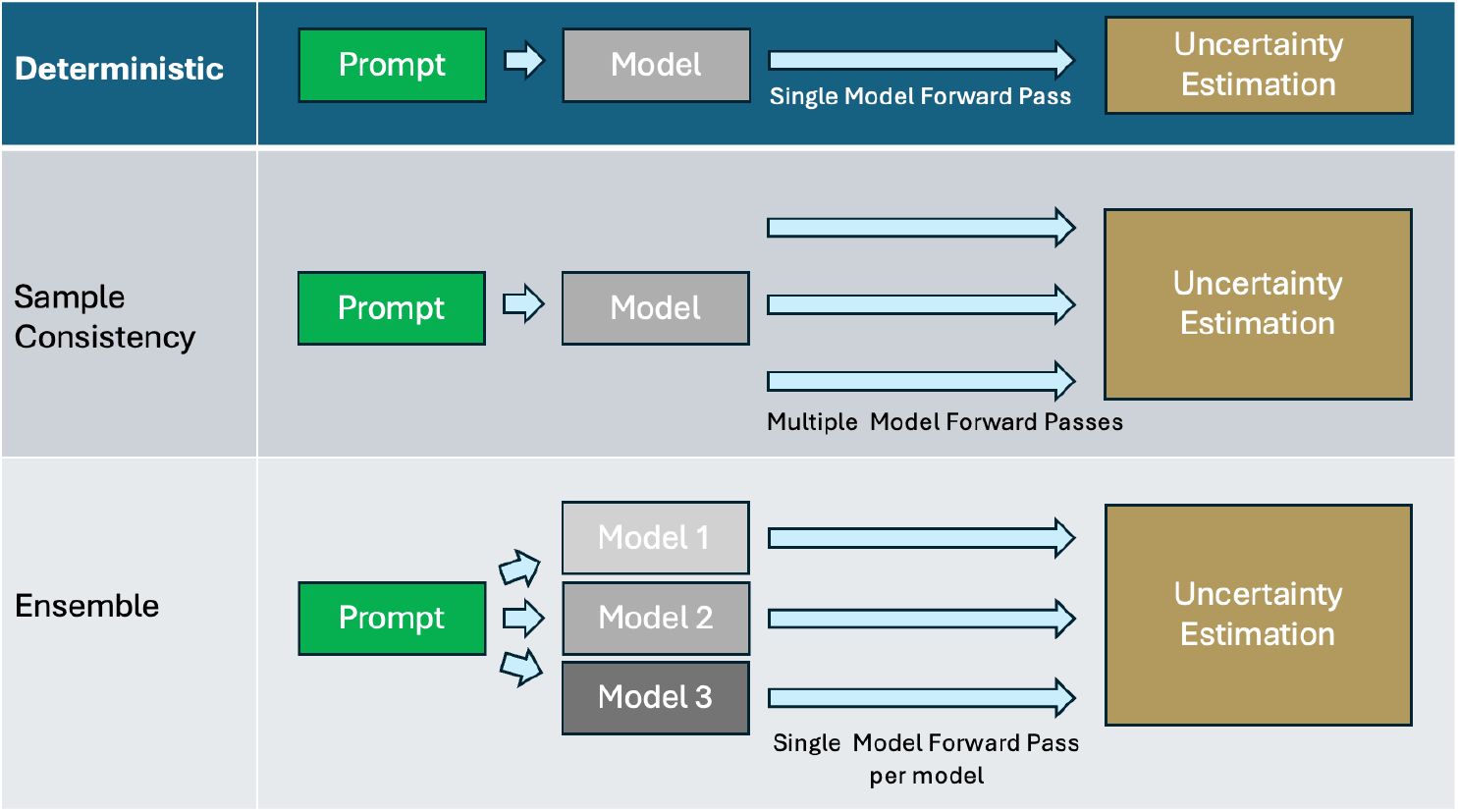
Illustration of the Deterministic, Consistency and Ensemble method families of estimating LLM uncertainty.

### Deterministic Methods

The two most common deterministic uncertainty measures that can be calculated from a single forward pass of a model are Confidence Elicitation^24^ and Token-Level Probabilities.^17^

Confidence Elicitation (CE) is the method of directly prompting the model to verbalize its degree of uncertainty on a scale from 0 to 100.^24^ Rivera et al^24^, Xiong et al^25^, and Zhou et al^26^ have evaluated Confidence Elicitation for the tasks of common sense reasoning, arithmetic reasoning, symbolic reasoning, and misinformation identification. Their investigations demonstrate that two-step Confidence Elicitation performs better than one-step Confidence Elicitation. Two-step Confidence Elicitation is the process of having a model generate a response to a question, and then resubmit the entire question-response to the model when performing Confidence Elicitation (see Figure 2).

**Figure 2.**
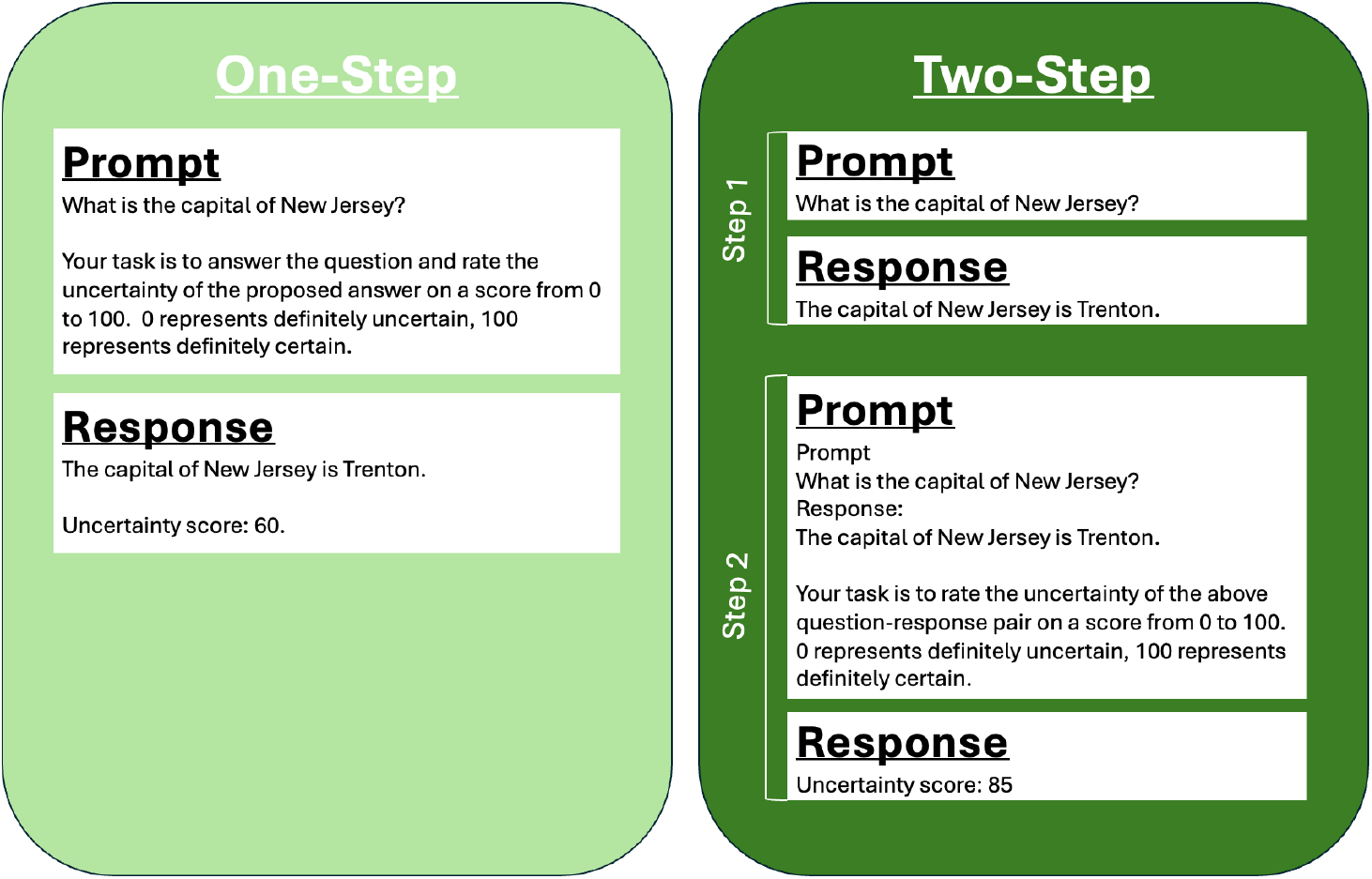
One-step Confidence Elicitation compared to two-step Confidence Elicitation.

Token-Level Probabilities (TLPs) are the other family of deterministic methods. TLP measures use generated token probabilities of an LLM response to calculate an uncertainty metric. Most commonly the uncertainty measure is calculated from the average, maximum, or minimum of generated token probabilities^17^, however more complex methods considering the semantic importance of each token have also been proposed.^27^ Figure 3 demonstrates how Token Level Probability measures are calculated from a model response.

**Figure 3.**
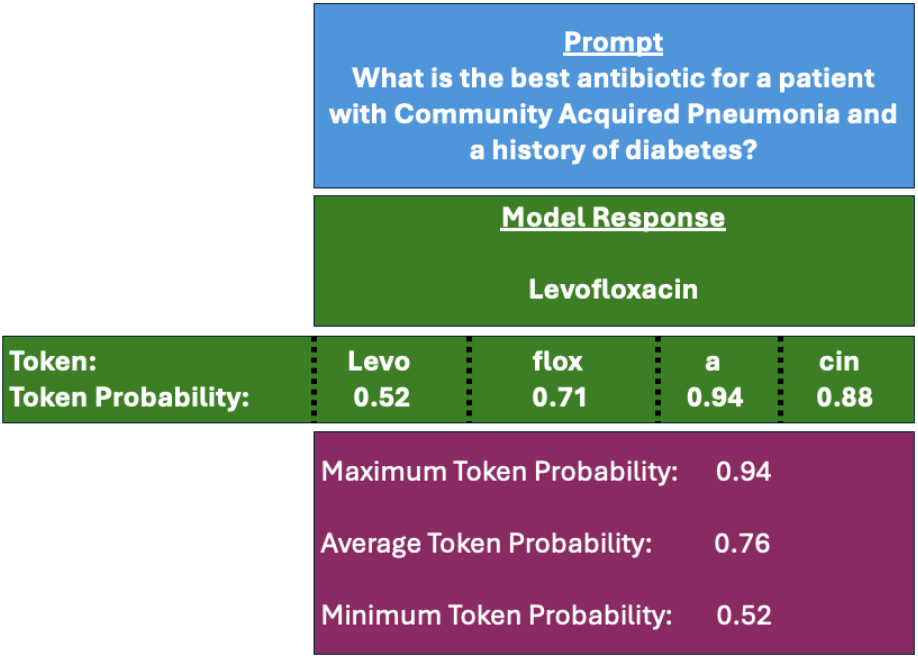
Example of Token Level Probabilities and how they can be leveraged to calculate uncertainty measures.

### Sample Consistency Methods

Sample Consistency (SC) methods leverage the stochastic behavior of LLMs to estimate uncertainty from inter-response agreement. Frequently Sample Consistency (also called Self-Consistency) methods are used to increase a model’s question-answering accuracy, but they can also be used to estimate uncertainty. The same prompt is run on a model multiple times and inter-response agreement (consistency) is used as a measure of uncertainty. The basic premise of SC methods is that a confident model will return similar answers with a high degree of agreement between answers, whereas an uncertain model will return drastically different answers (illustrated in Figure 4). Historically Sample Consistency methods were performed using Monte Carlo drop out, however given the size and complexity of LLMs along with their natural stochastic behavior, such methods are impractical and are not used.^28^

**Figure 4.**
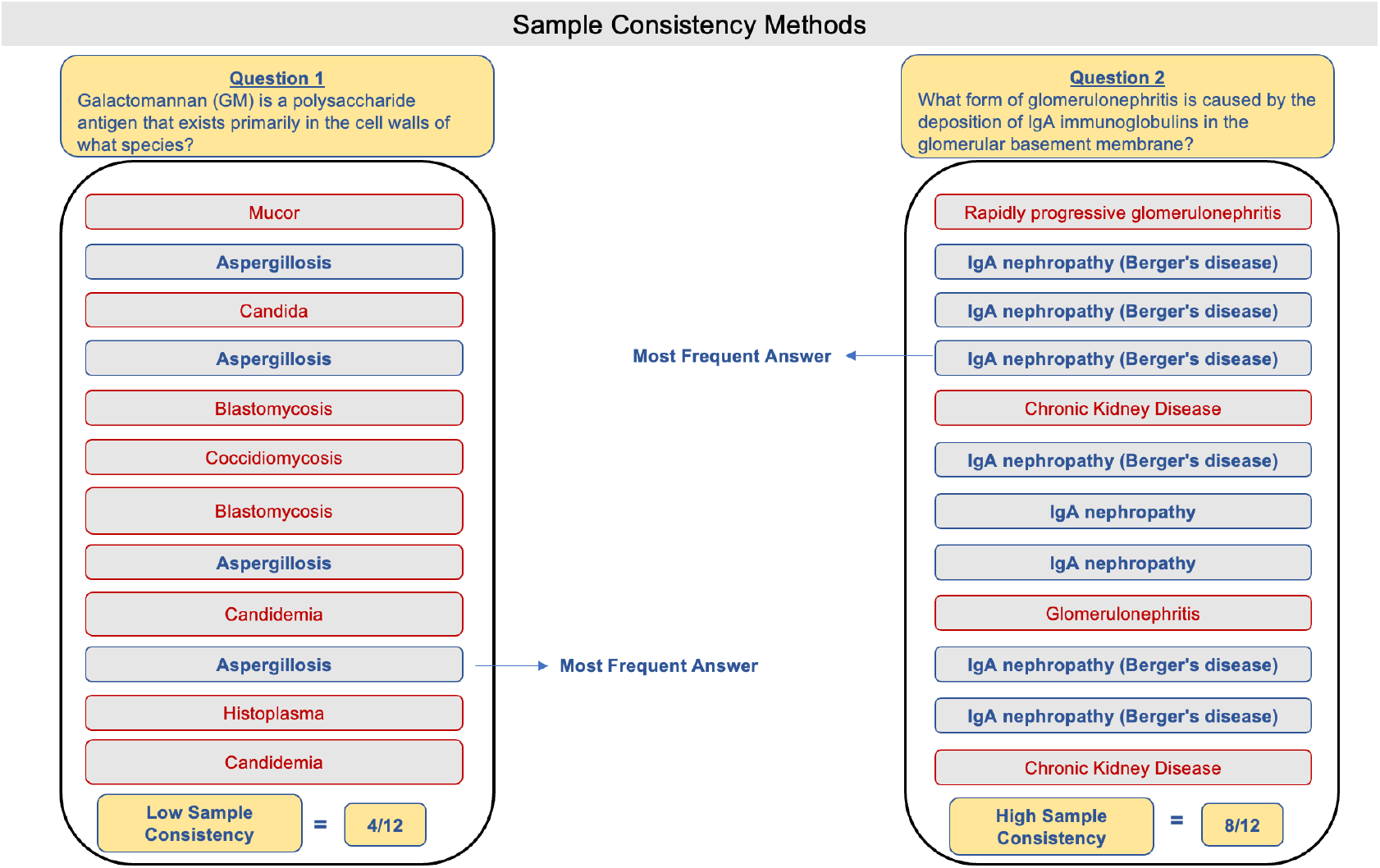
Demonstration of Sample Consistency (SC) methods. Multiple forward passes of a model for the same question are completed and the responses are compared for agreement. Higher similarity correlates to higher certainty.

Methods for measuring inter-response agreement include human annotation, cosine similarity between sentence embeddings, or BLEU score.^17,18^

## Methods

### Overview

In this study we evaluated three methods of uncertainty measurement for questions evaluating diagnosis and treatment selection clinical reasoning tasks. We evaluated Confidence Elicitation (CE), Token Level Probabilities (TLP) and Sample Consistency (SC) methods. Clinical reasoning questions were taken from three datasets: MedQA^29^, the New England Journal of Medicine Case Report Series^30^, and a custom dataset developed by authors JW and TS which we refer to as the Stanford dataset. The LLMs evaluated in this study are GPT 3.5, GPT 4, Llama2-70B, and Llama3-70B.

### Uncertainty Measures Evaluated

#### Two-step Confidence Elicitation

Two-step Confidence Elicitation (CE) in this investigation followed the methods outlined in Figure 2, where the model first generates a response to a question and then the entire question-response is resubmitted to perform Confidence Elicitation. We employed the same prompts as published by Zhou et al^31^ along with a system prompt encouraging the model to assume the role of an expert physician. Prompts can be found in supplemental information I. Full code can be found in Supplemental Information II and III.

#### Token Level Probabilities

Token Level Probabilities were used to calculate two uncertainty metrics: (1) average token probabilities of the LLM answer and (2) the minimum token probability of the tokens included in the LLM answer. Full code can be found in Supplemental Information II and III.

#### Sample Consistency

Sample Consistency methods was performed on 15 forward passes per question to assess inter-response agreement. A sample size of 15 was chosen because the investigation by Manakul et al^32^ demonstrated a plateau in Sample Consistency performance beyond a sample size of 15.

To assess inter-response agreement, we used two objective methods: GPT 4 Annotation and sentence embedding distance^17^ (Table 1). In GPT 4 Annotation, GPT 4 was asked to identify the most common response and tally the number of responses that agreed with the most common response. The tally acted as the inter-response agreement and corresponding uncertainty metric. The GPT 4 prompts used are provided in Supplemental Information IV.

**Table 1.** Overview of Sample Consistency measures of inter-response agreement.

| Sample Consistency Method | Final Answer Determination | Uncertainty Measure (Response Similarity Measure) |
| --- | --- | --- |
| GPT 4 Annotation | Most common response (via GPT 4 interpretation) | Fraction of responses with the most common response (via GPT 4 interpretation) |
| Sentence Embedding<br>a. e5-small-v2<br>b. S-PubMedBERT | Response with the embedding closest to the average. | Composite Cosine Similarity |

Sentence embedding distance was the other method used to assess inter-response agreement. Sentence embedding models convert LLM text responses into a numeric vector representation that summarizes the semantic meaning of the text response. These numeric vectors (embeddings) of an LLM response can then be compared to one another via quantitative metrics such as cosine similarity. In our experiment, we used sentence embeddings to determine the final answer as the response with the embedding closest to the average of all 15 sentence embeddings. Inter-response agreement was then determined by composite cosine similarity over all the sentence embeddings. Sentence embedding models e5-small-v2 and S-PubMedBERT were chosen because of their strong performance in the recent medical coding validation study by Excoffier et al.^33^ Full code can be found in Supplemental Information V.

### LLMs and Parameter Settings

The models evaluated in this study are GPT 3.5, GPT 4, Llama2 70B, and Llama3 70B, chosen to survey multiple generations of a popular closed source and open-source model. GPT 4 (gpt-4-0613) was accessed via API between March 7^th^ and March 23^rd^, 2024, GPT-3.5 (gpt-3.5-turbo-0125) was accessed via API between May 16^th^ and May 20^th^. Llama2 70B chat and Llama3 70B instruct were downloaded via huggingface^34^ and hosted on two A100 GPUs for question response and token probability generation, and then accessed via API^35^ for confidence elicitation.

LLM settings of top_p and top_k were kept at their respective baseline settings for each model. Temperature settings were trialed at three settings (0, 0.5 and 1.0), except for SC methods which were only trialed at settings of 0.5 and 1.0 because a temperature setting of 0 did not provide sufficient answer variability for inter-response comparison. Notably Llama temperature is unable to be set exactly at 0, so temperature was set at 0.01. Temperature settings higher than 1.0 produced disorganized responses on preliminary queries and therefore were not included in this investigation.

Seed for GPT 3.5 and GPT 4 prompts was set at 1, except for self-consistency where the 15 forward passes of the model had a seed corresponding to their index between 1 and 15.

### Datasets

The MedQA test set questions^29^ were screened to only include US Medical Licensing Exam Step 2 and 3 questions (Supplementary Information V). Only Step 2 and 3 questions were chosen because of their emphasis on evaluating general clinical reasoning and knowledge. The dataset totaled 418 questions, where 179 questions evaluated diagnostic clinical reasoning and 239 questions evaluated treatment selection clinical reasoning skills. Questions were modified to be open-ended rather than multiple choice to better simulate real clinical decision making and true calibration measurement.

The New England Journal of Medicine (NEJM) Case Report Series was used to test the task of diagnosis. The 200 most recent case reports before January 25^th^ 2024 were included in this dataset. The case scenario before expert interpretation was given to the model as input, and the expert’s diagnosis was used as the gold standard answer. Title and DOI information of case reports included can be found in Supplemental Information V.

The custom Stanford dataset consisted of 105 patient scenario questions which were developed to assess both diagnosis (54 questions) and treatment decision making (51 questions), modeled after USMLE step 2 and 3 board exam questions. The patient scenarios pertain to general medicine, inspired by challenging clinical cases encountered on general internal medicine wards by author TS, assessing practical knowledge of internal medicine. All patient-specific details have been changed and edited, retaining no original or identifying information. The dataset underwent quality control by two board certified internal medicine physicians.

Questions where the model generated an error response were not included in our analysis. The most common reasons a question was not included in the analysis were either the prompt exceeded the model’s context length or the LLM produced a response with incorrect formatting that prevented identification of a single articulated answer. The number of questions per dataset included in the final analysis can be found in Table 2 and full model responses can be found in Supplemental Information V.

**Table 2.**
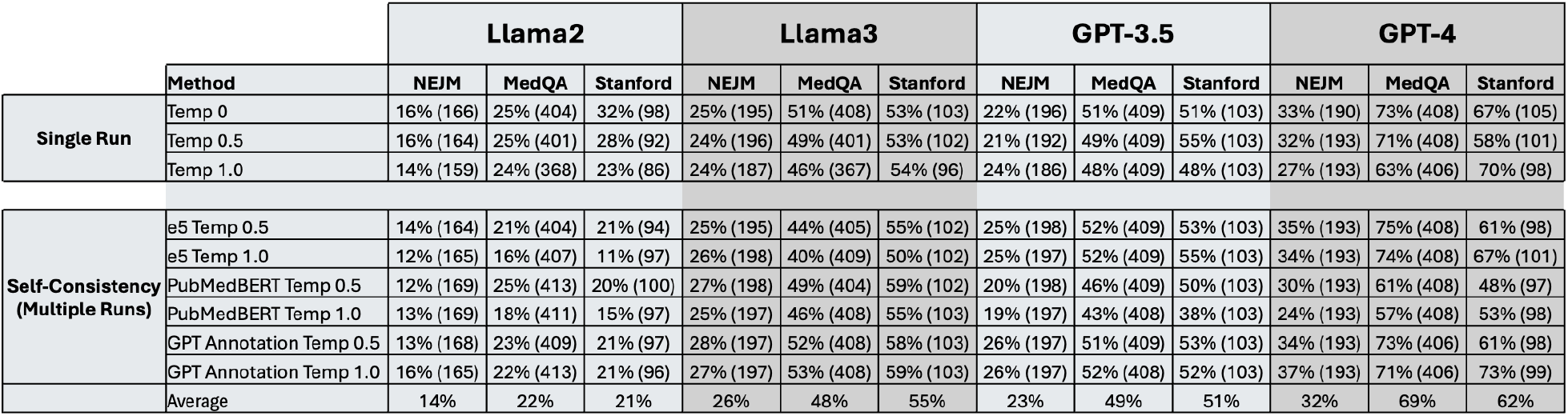
Accuracy of each language model for all datasets by method of model query. Accuracy is reported by a percentage, the number in parentheses is the number of questions answered and analyzed.

### Prompting

The prompts used for each dataset are provided in Supplemental Information V and are adapted from Savage et al.^36^ The prompts used chain-of-thought (CoT) and few shot strategies, except for the Llama2 NEJM analysis because the Llama2 token limit prevented use of these prompting strategies. Different few-shot examples were used for diagnosis and treatment selection questions.

### LLM Response Evaluation

LLM responses for the NEJM dataset were evaluated by three board certified Internal Medicine physicians, whereas the MedQA and Stanford datasets were evaluated by three MD/MBBS physicians. Each question was evaluated by two blinded evaluators and if there was disagreement in the grade assigned, a third evaluator determined the final grade. Any response that was felt to be equally correct and specific to the provided gold standard answer, was marked as correct. Physicians used UpToDate^37^, MKSAPP^38^, and StatPearls^39^ to verify accuracy of answers when needed. The graded files are provided in Supplemental Information V.

### Uncertainty Measure Statistical Evaluation

Discrimination was evaluated by logistic regression odds ratio and ROC AUC. An odds ratio p-value threshold of 0.0025 was used to reflect multiple hypotheses (multiple uncertainty measures) by Bonferroni Correction. ROC AUC confidence intervals (95%) were calculated using the sklearn python package with 4000 bootstrapped samples. An ROC AUC cut off of 0.7 was used to distinguish acceptable discriminative ability,^40^ and only uncertainty measures meeting this threshold were analyzed for calibration.

Calibration was evaluated by calibration plots as well as Expected Calibration Error and Brier Scores. Expected Calibration Error and Brier Scores were calculated using the same methods as described by Rivera et al^24^ with 10 bins. Statistical analysis code as well as Expected Calibration and Brier Score equations are included in Supplemental Information VIII.

## Results

Llama2 and Llama3 achieved average accuracies of 14% and 26% respectfully on the NEJM dataset, 22% and 48% for the curated MedQA dataset, and 21% and 55% on the Stanford dataset (Table 2). GPT 3.5 and GPT 4 achieved average accuracies of 23% and 32% respectfully on the NEJM dataset, 49% and 69% for the curated MedQA dataset, and 51% and 62% on the Stanford dataset (Table 2). Overall, the NEJM dataset seemed to be the most challenging, while the MedQA and Stanford were less difficult. Our accuracies for the MedQA and NEJM datasets are different than other reported investigations using the same datasets^41–44^ because our study used open-ended questions instead of multiple choice questions and used only a subset of the MedQA test set questions that evaluate diagnostic and treatment selection clinical reasoning skills at a Step 2 and 3 level rather than the entire test set.

Interrater agreement was 93% over all datasets, with a Cohen Kappa Statistic 0.85 representing strong agreement amongst graders. Full grades can be found in Supplemental Information V.

### Discrimination

Evaluation of each uncertainty method’s ability to discriminate between correct and incorrect answers was first investigated using odds ratios (Figure 5.). Across all models except Llama2, Sample Consistency methods showed statistically significant discrimination. Token Level Probability measures showed statistical significance inconsistently, more frequently with the GPT model family rather than the Llama model family. Confidence Elicitation methods showed less discriminative ability with early model versions (GPT 3.5 and Llama2) but demonstrated relatively consistent statistical significance with newer model versions (GPT 4 and Llama3).

**Figure 5.**
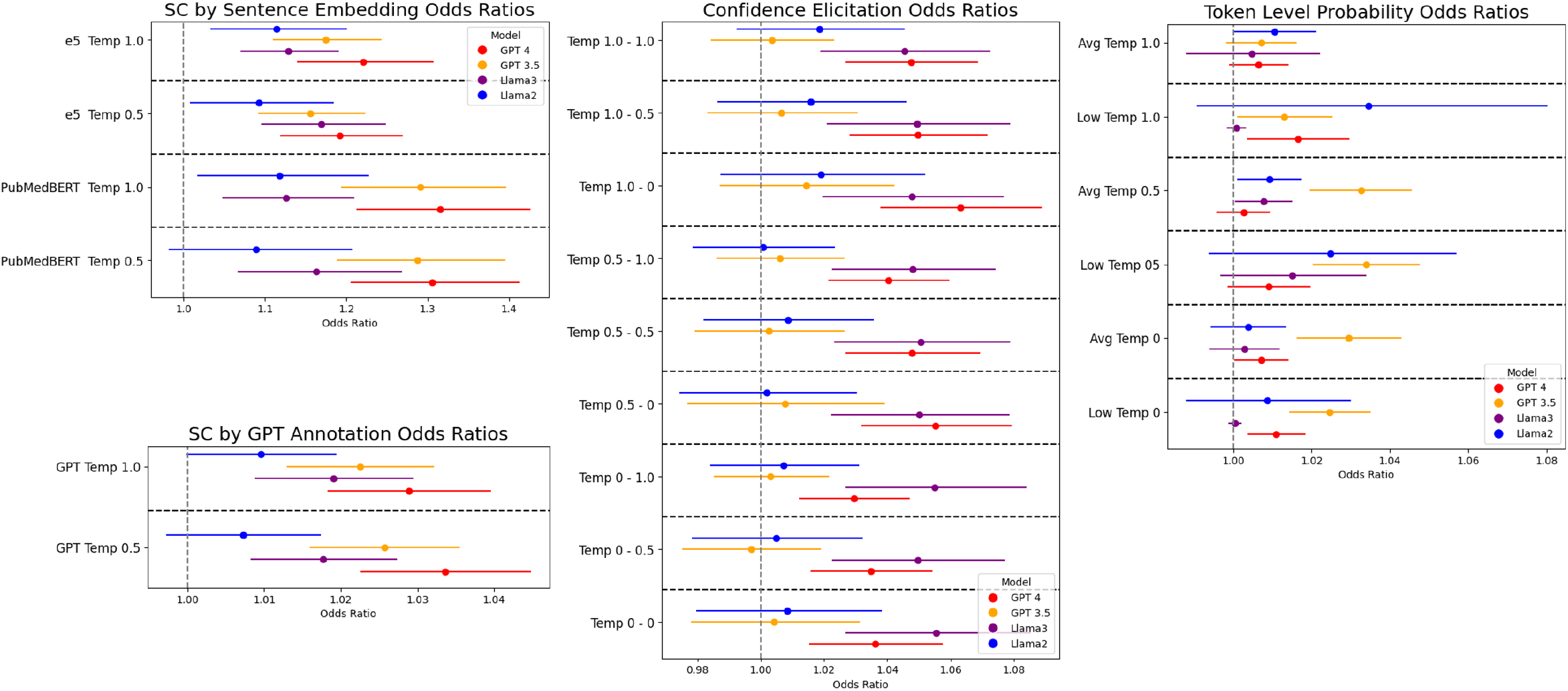
Forest plot of Odds ratio (OR) and Confidence Interval with a p value threshold of 0.0025 to account for multiple hypothesis testing. Methods whose Confidence Interval does not intersect with the dotted line corresponding to an odds ratio of 1.0 reach statistical significance. Odds Ratios were calculated using all questions (combining diagnosis and treatment selection subsets). Sample Consistency is abbreviated SC.

Discrimination was also evaluated using ROC AUC, finding a few uncertainty measures reached our ROC AUC threshold of 0.7 (Figure 6 and 7). Overall Sample Consistency methods generally outperformed Token Level Probability and Confidence Elicitation methods for both diagnosis and treatment selection tasks, with the exceptions of Llama3 for diagnosis (Confidence Elicitation) and Llama2 for treatment selection (Token Level Probabilities). Top-performing Sample Consistency methods achieved ROC AUC values for diagnosis and treatment selection tasks respectively of 0.77 and 0.68 for GPT 3.5, 0.79 and 0.71 for GPT 4, 0.77 and 0.56 for Llama2, and 0.68 and 0.75 for Llama3.

**Figure 6.**
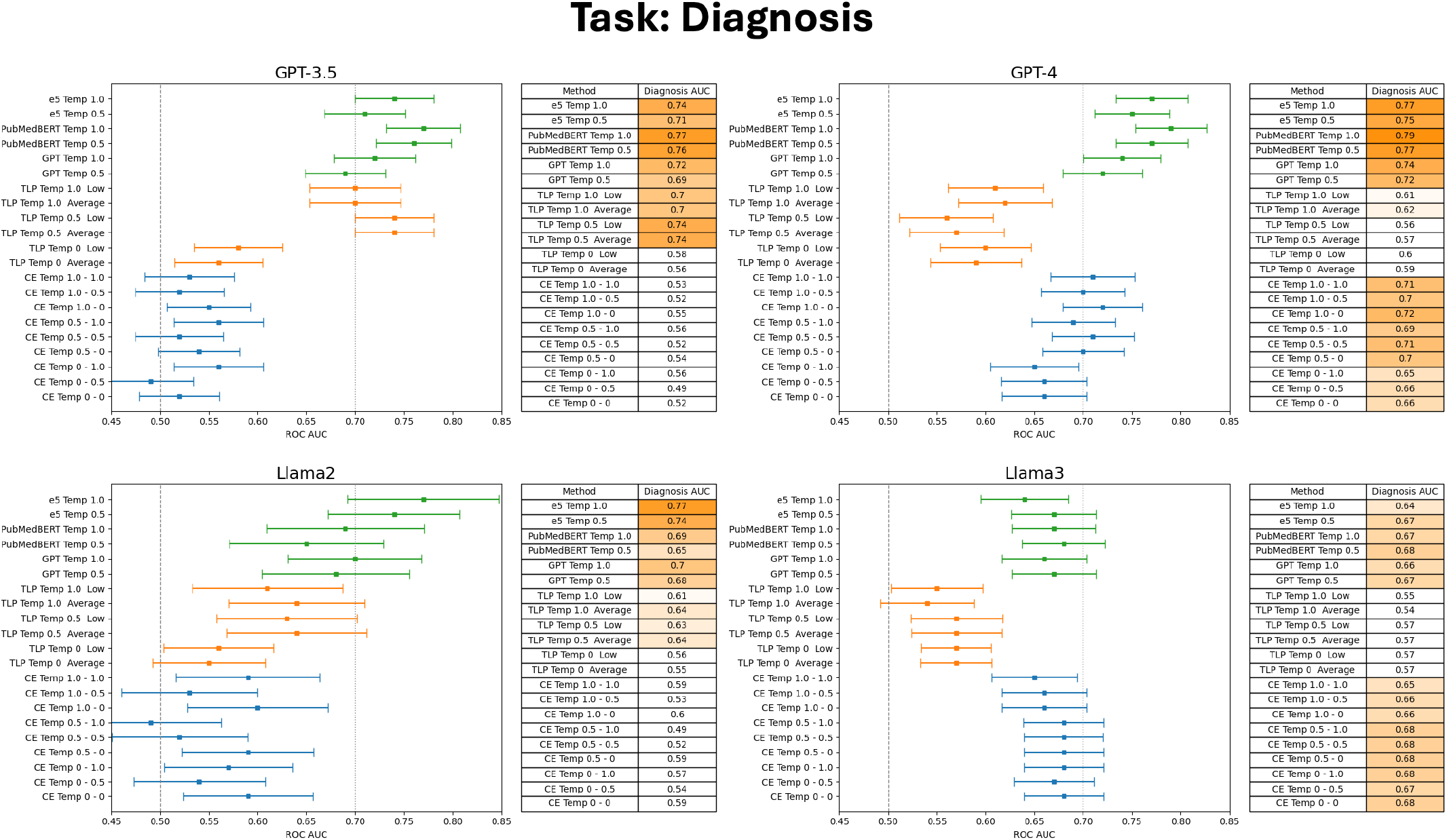
Receiver Operator Characteristic Area Under the Curve with 95% Confidence Intervals for questions assessing diagnostic clinical reasoning. Sample Consistency methods are colored green, Token Level Probability (TLP) methods are colored orange, and Confidence Elicitation (CE) methods are colored blue. A higher ROC AUC corresponds to better discrimination by the uncertainty metric.

**Figure 7.**
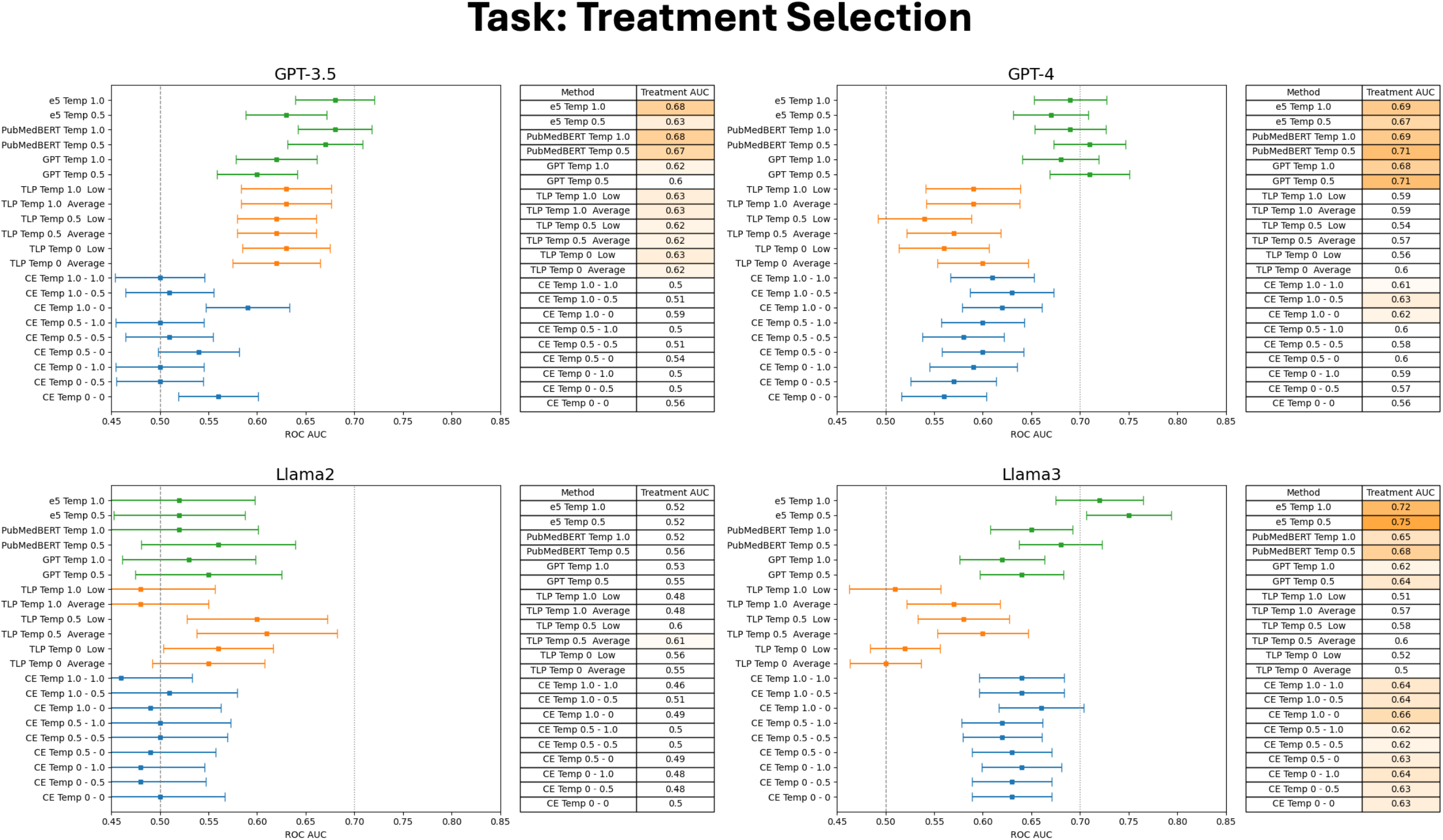
Receiver Operator Characteristic Area Under the Curve with 95% Confidence Intervals for questions assessing treatment selection reasoning. Sample Consistency methods are colored green, Token Level Probability (TLP) methods are colored orange, and Confidence Elicitation (CE) methods are colored blue. A higher ROC AUC corresponds to better discrimination by the uncertainty metric.

Early generation models (GPT 3.5 and Llama2) demonstrated better discriminative ability by ROC AUC with Token Level Probability methods rather than Confidence Elicitation. In contrast later generation models (GPT 4 and Llama3) had worse Token Level Probability discrimination and better performance with Confidence Elicitation. No change in discriminative performance was observed between different temperature settings within each uncertainty metric family.

Discriminative performance for diagnosis tasks was higher than for treatment selection tasks (Figures 6 and 7). This trend was observed for nearly all uncertainty metrics except Sample Consistency by e5 sentence embedding with Llama3.

### Calibration

Calibration analysis was performed for uncertainty measures that met our discrimination ROC AUC threshold (0.7) for the task of diagnosis. Calibration plots in Figure 8 show GPT Annotation showed the most accurate calibration amongst Sample Consistency methods, most closely approximating perfect calibration (represented by the black dashed line). In contrast Sample Consistency methods that used Sentence Embeddings demonstrated poor calibration due to their limited range of values (Figure 8). Composite cosine similarity values clustered between 0.8 and 1.0, significantly limiting the calibration of embedding-based measures. This limitation is reflected in their respective Expected Calibration Error (ECE) and Brier’s scores, where GPT Annotation yielded lower (better) scores for both metrics (Figure 9).

**Figure 8.**
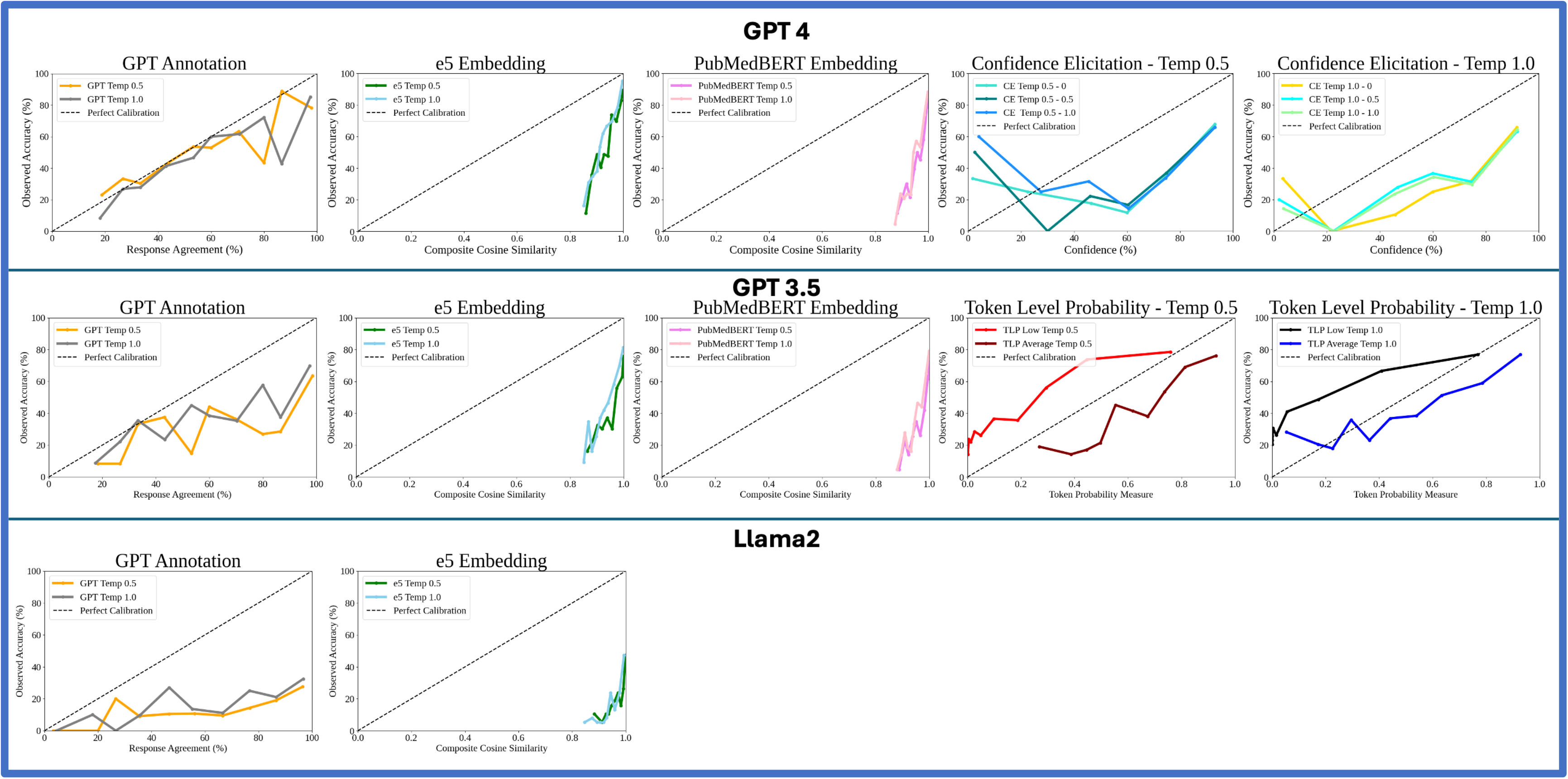
Calibration plots for uncertainty metrics with an ROC AUC greater than 0.7 for diagnostic questions. Calibration plots compare the model’s confidence metric against the observed accuracy. Metrics who closely align with the observed accuracy are highly useful to physicians when making clinical decisions. Perfect calibration is denoted by the dashed black line. Notably Sentence Embedding and Confidence Elicitation metrics demonstrate miscalibration and overconfidence.

**Figure 9.**
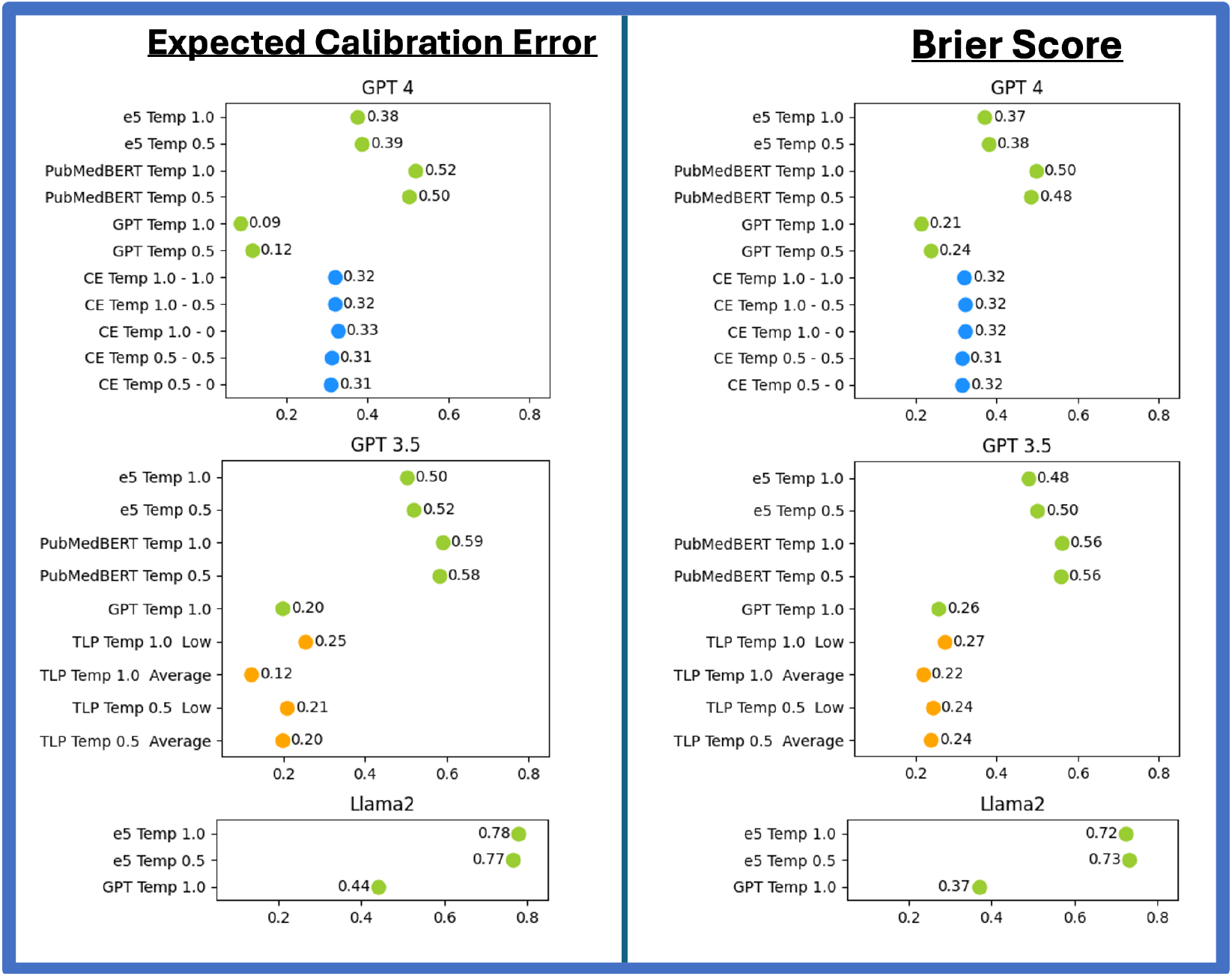
Calibration measures of Expected Calibration Error and Brier Scores for uncertainty metrics that met our discrimination ROC AUC threshold for the task of diagnosis. A lower score closer to zero indicates more accurate calibration. Sample Consistency methods are colored green, Token Level Probability methods are colored orange, and Confidence Elicitation methods are colored blue.

Token Level Probability measures reached our ROC AUC threshold with GPT 3.5 at temperature settings of 0.5 and 1.0, and were included in our calibration analysis. Both Token Level Probability average and minimum demonstrated accurate calibration, with similar Expected Calibration Error and Brier’s scores to Sample Consistency GPT Annotation (Figures 8 and 9).

Confidence Elicitation measures reached our ROC AUC threshold with GPT 4 at temperature settings of 0.5 and 1.0, and were also included in our calibration analysis. Confidence Elicitation measures demonstrated more accurate calibration compared to Sample Consistency Sentence Embedding methods (e5 and PubMedBERT), but had less accurate calibration than GPT Annotation by both Expected Calibration Error and Brier score (Figures 9).

## Discussion

The results of our investigation show Sample Consistency (SC) methods largely have the best discriminative ability amongst the uncertainty metrics evaluated for the medical tasks of diagnosis and treatment selection. Token Level Probability and Confidence Elicitation methods generally demonstrated worse discrimination, with ROC AUC values frequently below our threshold of 0.7, rendering them less clinically useful. Therefore our results suggest Sample Consistency methods should be the primary uncertainty measures used for medical applications.

Amongst the Sample Consistency methods evaluated, GPT Annotation had the most accurate calibration. Uncertainty prediction via Sample Consistency with GPT Annotation correlated well with observed accuracy, a valuable attribute that provides the clinician a tangible and actionable estimate of the model’s uncertainty. In contrast Sample Consistency Sentence Embedding methods (e5 and PubMedBERT) demonstrated high discrimination but poor calibration due to their clustering of values between 0.8 and 1.0 (Figure 8). This irregular range of values did not correlate well with observed accuracy and makes Sample Consistency Sentence Embedding methods less actionable for clinicians. Therefore, unless the user has a large number of reference cases available to re-calibrate a metric via binning, GPT Annotation is the optimal Sample Consistency method that balances strong discrimination with accurate calibration.

The limitations of Sample Consistency methods as an uncertainty metric are computational cost and time. Sample Consistency methods require multiple times more compute because the model must be run many times to calculate inter-response agreement. These limitations make Sample Consistency methods a poor choice for tasks that are time sensitive or occur frequently. Though the discriminative ability of Sample Consistency methods are impressive, their superior performance must be weighed against their resource requirements.

Discrimination results stratified by clinical reasoning task (Figures 6 and 7) found most uncertainty metrics studied had better discriminative ability for diagnosis questions compared to treatment questions. This finding identifies a potential limitation of current LLM models and uncertainty metrics, that they may be ill-equipped to assess treatment selection scenarios. This deficit may be due to limitations in language model training data, where text detailing diagnostics is more prevalent than text discussing treatment selection. Alternatively, this finding could be explained by the nature of treatment selection questions, where there are often many semi-correct treatments options but only one “best” choice. The presence of multiple semi-correct treatments may limit the model’s internal mechanisms to identify whether a suggested treatment is ultimately correct or incorrect. Regardless of the mechanism, our findings suggest caution when using uncertainty measures for treatment selection tasks.

Our investigation also validates concerns within the medical community that LLMs are overconfident when answering medical questions. Calibration plots shown in Figure 8 as well as Supplemental Information VII show Confidence Elicitation methods are nearly universally overconfident. Therefore we advise physicians to be cognizant that LLM frequently state responses with overconfidence, especially when using simple uncertainty measures such as Confidence Elicitation, and to use caution when integrating LLMs into patient care workflows.

The strengths of our investigation are a robust investigation of uncertainty metrics over four LLMs and three datasets. We also use free response questions where previous studies have been limited to multiple choice questions.^3,17,23,45^ This allowed for a unique evaluation of calibration measurement that more closely simulated clinical practice.

A limitation of our study is that our investigation was limited to only US-centric question sets and the English language, therefore we cannot generalize our findings to non-English languages and non-US centric question sets. Furthermore, we cannot generalize our findings for models that are fine-tuned on medical text. We hope that future studies can iterate on our proposed metrics and use our open dataset as a benchmark for additional evaluation.

## Conclusions

Sample Consistency methods have superior discrimination as a measure of LLM uncertainty compared to Token Level Probabilities and Confidence Elicitation for clinical reasoning tasks of diagnosis and treatment selection. Sample Consistency by Sentence Embedding Cosine Similarity yields the best discrimination of the measures evaluated, while Sample Consistency by GPT Annotation has better balance between discrimination and accurate calibration. Medical providers should exercise caution when asking LLMs to verbalize their confidence (Confidence Elicitation methods), as LLMs are consistently over-confident in their apparent assertions.

## Data Availability

All data is available for review at: https://doi.org/10.6084/m9.figshare.25962529.v1

## Supplemental Information I

Two-step Calibration Elicitation prompts, adapted from Zhou et al^31^:

### System Prompt

You are an expert physician, asked to rate the uncertainty of an answer.

### Prompt

** Question Prompt **

** LLM Provided Answer **

Your task is to rate the uncertainty of the proposed answer on a score from 0 to 100, where 0 represents definitely uncertain and 100 represents definitely certain. Please, only answer with your score.

## Supplemental Information II

Generation code for GPT 3.5 and GPT-4. File available on figshare: https://doi.org/10.6084/m9.figshare.25962529.v1.

## Supplemental Information III

Generation code for Llama-2 and Llama-3. File available on figshare: https://doi.org/10.6084/m9.figshare.25962529.v1.

## Supplemental Information IV

Prompts used in this study:

### Base Prompt used for the MedQA and Stanford dataset Diagnosis Questions

Answer the diagnosis or management question below. First we provide two examples. Write your answer in brackets (ex. [Answer]).

#### Example Question 1

Shortly after undergoing a bipolar prosthesis for a displaced femoral neck fracture of the left hip acquired after a fall the day before, an 80-year-old woman suddenly develops dyspnea. The surgery under general anesthesia with sevoflurane was uneventful, lasting 98 minutes, during which the patient maintained oxygen saturation readings of 100% on 8 L of oxygen. She has a history of hypertension, osteoporosis, and osteoarthritis of her right knee. Her medications include ramipril, naproxen, ranitidine, and a multivitamin. She appears cyanotic, drowsy, and is oriented only to person. Her temperature is 38.6°C (101.5°F), pulse is 135/minute, respirations are 36/min, and blood pressure is 155/95 mm Hg. Pulse oximetry on room air shows an oxygen saturation of 81%. There are several scattered petechiae on the anterior chest wall. Laboratory studies show a hemoglobin concentration of 10.5 g/dL, a leukocyte count of 9,000/mm3, a platelet count of 145,000/mm3, and a creatine kinase of 190 U/L. An ECG shows sinus tachycardia. What is the most likely diagnosis?

#### Rationale

The patient had a surgical repair of a displaced femoral neck fracture. The patient has petechiae. The patient has a new oxygen requirement, meaning they are having difficulty with their breathing. This patient most likely has a fat embolism.

Answer: [Fat Embolism]

#### Example Question 2

A 55-year-old man comes to the emergency department because of a dry cough and severe chest pain beginning that morning. Two months ago, he was diagnosed with inferior wall myocardial infarction and was treated with stent implantation of the right coronary artery. He has a history of hypertension and hypercholesterolemia. His medications include aspirin, clopidogrel, atorvastatin, and enalapril. His temperature is 38.5Â°C (101.3°F), pulse is 92/min, respirations are 22/min, and blood pressure is 130/80 mm Hg. Cardiac examination shows a high-pitched scratching sound best heard while sitting upright and during expiration. The remainder of the examination shows no abnormalities. An ECG shows diffuse ST elevations. Serum studies show a troponin I of 0.005 ng/mL (N < 0.01). What is the most likely cause of this patient’s symptoms?

#### Rational

This patient is having chest pain. They recently had a heart attack and has new chest pain, suggesting he may have a problem with his heart. The EKG has diffuse ST elevations and he has a scratching murmur. This patient likely has Dressler Syndrome.

Answer: [Dressler Syndrome]

XXXXXXXXX

Real Question:

### Base Prompt used for the MedQA and Stanford dataset Treatment Selection Questions

Answer the diagnosis or management question below. First we provide two examples. Write your answer in brackets (ex. [Answer]).

#### Example Question 1

A 39-year-old woman is brought to the emergency room by her fiance for severe abdominal pain for the past 5 hours. She was watching TV after dinner when she felt a sudden, sharp, 10/10 pain at the epigastric region that did not go away. Ibuprofen also did not help. She reports recurrent abdominal pain that would self-resolve in the past but states that, Äúthis one is way worse.,Äù Her past medical hi story is significant for diabetes and an appendectomy 2 years ago. The patient endorses nausea and 1 episode of emesis, but denies fevers, chills, chest pain, shortness of breath, diarrhea, constipation, urinary symptoms, paresthesia, or weakness. She used to smoke marijuana in college and drinks about 2 beers a week. A physical examination demonstrates an overweight woman in acute distress with diffuse abdominal tenderness. Her vitals are within normal limits. Laboratory values are shown below:

Hemoglobin: 12 g/dL

Hematocrit: 34%

Leukocyte count: 4,900/mm^3 with normal differential

Platelet count: 160,000/mm^3

Serum:

Na+: 138 mEq/L

Cl-: 98 mEq/L

K+: 4.8 mEq/L

HCO3-: 25 mEq/L

Glucose: 123 mg/dL

Ca2+: 6.9 mg/dL

AST: 387 U/L

ALT: 297 U/L

ALP: 168 U/L

Lipase: 650 U/L (Normal 0, Äì 160 U/L)

What is the best next step in the workup of this patient?

##### Rationale

This patient is presenting with epigastric pain as well as transaminitis and elevated lipase. These findings are concerning for choledocholithiasis, cholangitis and gallstone pancreatitis. Given her absence of fever and chills the most likely diagnoses are gallstone pancreatitis and choledocholithiasis. To better evaluate these diagnoses, a right upper quadrant ultrasound would be most appropriate.

Answer: [Right Upper Quadrant Ultrasound]

#### Example Question 2

A 54-year-old woman presents to her gynecologist complaining of incontinence. She reports leakage of a small amount of urine when she coughs or laughs as well as occasionally when she is exercising. She denies any pain with urination. She underwent menopause 2 years ago and noted that this problem has increased in frequency since that time. Her history is significant only for three uncomplicated pregnancies with vaginal births. Urinalysis, post-void residual, and cystometrogram are conducted a nd all show normal results. The patient’s vital signs are as follows: T 37.5 C, HR 80, BP 128/67, RR 12, and SpO2 99%. Physical examination is significant for pelvic organ prolapse on pelvic exam. what is a reasonable first step in the management of this patient’s condition?

##### Rationale

This patient with a history of multiple vaginal births and incontinence presents with leakage of urine when laughing. Her gynecologic history and described symptoms are consistent with stress incontinence. The most appropriate treatment for stress incontinence would be to first teach the patient how to perform kegel exercises.

Answer: [Kegel Exercises]

### Base GPT-4 Prompt used for the NEJM datasets

Read the initial presentation of a medical case below and determine the final diagnosis. Assume that all of the relevant details from figures and tables have been explained in the text. When providing your rationale, USE STEP-BY-STEP DEDUCTION TO IDENTIFY THE CORRECT RESPONSE. After you provide your rationale, provide a single, specific diagnosis for the case in less than 10 words. Clearly label your Rationale in square brackets and then your answer in asterisks.

Below we provide an example:

Example Case: DOI: 10.1056/NEJMcpc1413303

[Rationale]

This patient has oral ulcers, which can be associated with autoimmune and infectious diseases. The patient has a negative infectious work up and did not respond to antibiotics, which supports an autoimmune process. The patient has genital ulcers, which are associated with the autoimmune process of Behcets disease. Nodules of the legs further support an autoimmune process such as Behcets disease. Symmetric arthralgias are seen in a majority of patients with Behcets disease. Fever can be seen in systemic autoimmune processes such as Behcets disease. The rash is described as pustular, which can be seen in pathergy phenomenon, a highly specific sign for Behcets disease.

*Behcets Disease*

==========

Real case:

### Base Llama-2 Prompt used for the NEJM datasets

Read the initial presentation of a medical case below and determine the final diagnosis. Assume that all of the relevant details from figures and tables have been explained in the text. Provide a single, specific diagnosis for the case in less than 10 words. Clearly label your Answer in square brackets (ex. [Diagnosis: Cellulitis]).

===========

Case:

### Experimental Prompts

1. Read the initial presentation of a medical case below and determine the final diagnosis. Assume that all of the relevant details from figures and tables have been explained in the text. When providing your rationale, USE STEP-BY-STEP DEDUCTION TO IDENTIFY THE CORRECT RESPONSE. After you provide your rationale, provide a single, specific diagnosis for the case in less than 10 words. Example Case: DOI: 10.1056/NEJMcpc1413303 Rationale (REMEMBER TO USE STEP BY STEP DEDUCTION): This patient has oral ulcers, which can be associated with autoimmune and infectious diseases. The patient has a negative infectious work up and did not respond to antibiotics, which supports an autoimmune process. The patient has genital ulcers, which are associated with the autoimmune process of Behcet,Äö√Ñ√¥s disease. Nodules of the legs further support an autoimmune process such as Behcet,Äö√Ñ√¥s disease. Symmetric arthralgias are seen in a majority of patients with Behcet,Äö√Ñ√¥s disease. Fever can be seen in systemic autoimmune processes such as Behcet,Äö√Ñ√¥s disease. The rash is described as pustular, which can be seen in pathergy phenomenon, a highly specific sign for Behcet,Äö√Ñ√¥s disease. Diagnosis: Behcet,Äö√Ñ√¥s disease is the most likely diagnosis. === Case: Rationale (REMEMBER TO USE STEP BY STEP DEDUCTION): Diagnosis:
2. Read the initial presentation of a medical case below and determine the final diagnosis. Assume that all of the relevant details from figures and tables have been explained in the text. When providing your rationale, USE STEP-BY-STEP DEDUCTION TO IDENTIFY THE CORRECT RESPONSE. After you provide your rationale, provide a single, specific diagnosis for the case in less than 10 words. Clearly label your Rationale in square brackets and then your answer in asterisks. [Rationale] *Answer* Real case:

## Supplemental Information V

Graded csv files.

Files available on figshare: https://doi.org/10.6084/m9.figshare.25962529.v1.

## Supplemental Information VI

Uncertainty Measure Generation Code Llama Models.

File available on figshare: https://doi.org/10.6084/m9.figshare.25962529.v1.

## Supplemental Information VII

Uncertainty Measure Generation Code GPT Models.

File available on figshare: https://doi.org/10.6084/m9.figshare.25962529.v1.

## Supplemental Information VIII

Statistical Calculations Code

File available on figshare: https://doi.org/10.6084/m9.figshare.25962529.v1.

Expected Calibration Error

m = number of bins

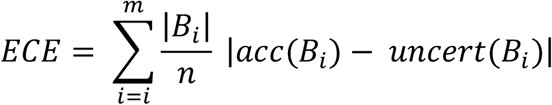

Brier Score

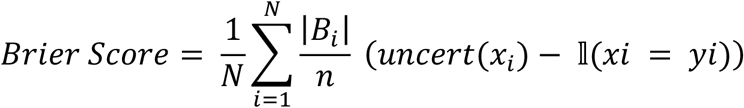

## Supplemental Information IX

Discriminative ROC AUC sub-analyses stratified by dataset and clinical reasoning task.

Files available on figshare: https://doi.org/10.6084/m9.figshare.25962529.v1.

## Supplemental Information X

Diagnosis task calibration plots for all Confidence Elicitation settings and models, demonstrating how large language models are frequently overconfident when answering medical questions.

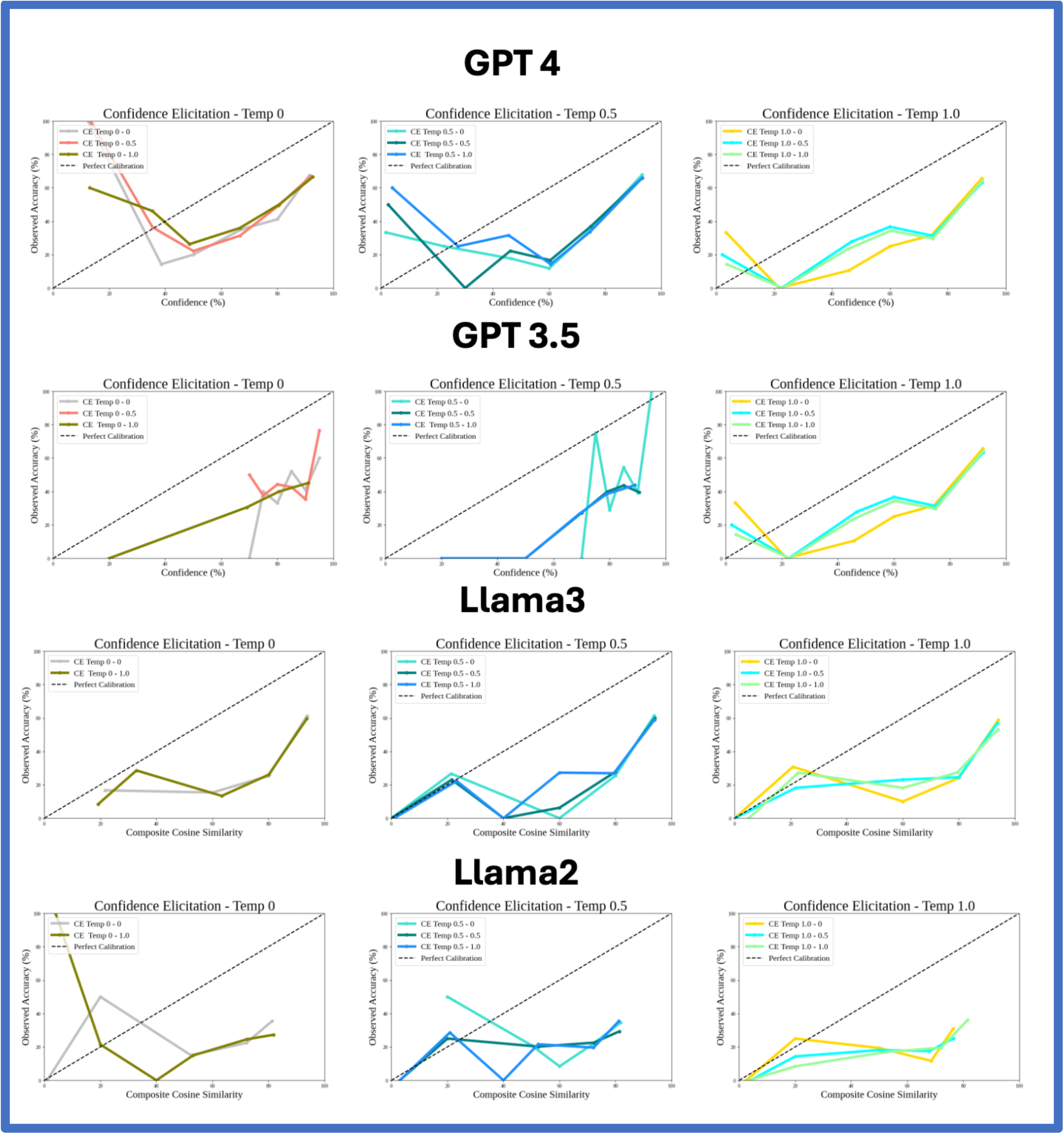

